# Prevalence of functional dyspepsia in medical students: protocol for a systematic review and meta-analysis

**DOI:** 10.1101/2020.12.17.20248361

**Authors:** Giulior A. Marrull Caldas, Nicolas Silva Farfán, Andres Penny Sauca, Steffi N. Roca Ortega, Jessica Hanae Zafra-Tanaka

**Author notes:** **Correspondence:** Steffi N. Roca Ortega.

## Abstract

**Background:** Dyspepsia is defined as a syndrome that comprises a series of chronic and recurrent disorders in the upper digestive tract. According to a meta-analysis carried out in 2014, the worldwide prevalence of dyspepsia is 20.8%, being the highest prevalence found in South America with 37.7%. It was found that there are populations that present a higher prevalence of dyspepsia that is associated with high levels of stress, such as those that can be found in medical students, so It would be important to evaluate the population of medical students, since the medical career, by its nature, represents a great academic load on the student who may later suffer from gastrointestinal diseases.

**Review question:** What is the prevalence of functional dyspepsia in medical students?

**Objective:** To determine the prevalence of dyspepsia in medical students and to assess genre differences

**Methods:** We will conduct a systematic review and meta-analysis. We will use the following search engines: PubMed, Global Index Medicus and EMBASE using search terms related to functional dyspepsia and medical students. We will include observational studies (either cross-sectional or cohort studies) that assessed the prevalence of functional dyspepsia among this particular population.

**Conclusion:** Currently, no systematic review or meta-analysis has been conducted about the prevalence of functional dyspepsia among this population of students. The aim of this work is to summarize the available literature regarding this topic.

## Background

Dyspepsia is defined as a syndrome that comprises a series of chronic and recurrent disorders in the upper digestive tract. It is considered a multifactorial disease, whose main symptoms include the sensation of early fullness, belching, epigastric discomfort and nausea, among others. (1) Dyspepsia can be classified into 3 categories: a) organic dyspepsia, in which the organic or metabolic cause of the disorder was found, b) functional dyspepsia, in which dyspepsia has already been investigated, but no apparent cause has been found, and c) uninvestigated dyspepsia.

The frequency of the disease varies according to the study population and the criteria used for its classification. Most of the studies have been conducted in Northern Europe and Southeast Asia. According to a meta-analysis carried out in 2014, the worldwide prevalence of dyspepsia is 20.8%, being the highest prevalence found in South America with 37.7%. In this same study, it was found that the prevalence of uninvestigated dyspepsia in Peru was between 25 - 29.9%, using epigastric or upper abdominal pain as diagnostic criteria. (2)

In a multicenter study carried out in Latin America, it was found that there are populations that present a higher prevalence of dyspepsia that is associated with high levels of stress, such as those that can be found in medical students. It would be important to evaluate the population of medical students, since the medical career, by its nature, represents a great academic load on the student who may later suffer from gastrointestinal diseases. (1)

Among pre-medical and medical students, there are several studies using Rome III criteria for establishing prevalence of functional dyspepsia (FD). A study regarding the overlap of FD and irritable bowel syndrome (IBS) in Peru found a prevalence of 16.9% of FD using these criteria among medical students in a private university. (3) Likewise, a multicentric study in Latin-American universities, which investigated associated risk factors for uninvestigated dyspepsia, concluded that the prevalence among students from four Latin-American faculties of medicine is higher than the reported for the general population in South America, nearly 46%. (1)

So far, no previous systematic reviews have been found for the prevalence of dyspepsia among this particular population of students. Taking into account primary studies that report a higher prevalence of dyspepsia among medical students over the general population, it would be noteworthy to summarize these findings. The objective of this study is to determine the prevalence of dyspepsia among medical students.

## Methods

### Study design

This systematic review and meta-analysis will be conducted according to the preferred reporting items for systematic reviews and meta-analyses (PRISMA guidelines).

### Eligibility criteria

1. Participants: Studies including medical students for the determination of uninvestigated/functional dyspepsia prevalence.
2. Exposure: Studying medicine
3. Outcome: Prevalence of uninvestigated/functional dyspepsia in the general population.
4. Types of studies: This study will include observational studies (either cross-sectional or cohort studies).

### Exclusion criteria

- The following study designs: case control, case reports, case series, letters to the editor, editorial, narrative review, systematic reviews, correspondence, short communications, technical notes, commentaries and pictorial essays.
- We will exclude studies that include prevalence for organic dyspepsia.
- Studies including students from faculties other than medicine.
- We will only include studies that have an English or Spanish translation.

### Literature Search and Data collection

- These databases will be used to start the search: 1) PubMed, 2) Global Index Medicus and 3) EMBASE.
- Two authors will independently screen titles and abstracts and select those which meet the eligibility criteria. Then two authors will independently review full-texts select studies for inclusion in the review. Inconsistencies will be discussed with a third reviewer to get to a consensus.
- The references used in the studies included in this work will also be reviewed in order to find other potential articles to include.
- The authors will record the results of included studies and show them on a PRISMA flow diagram.
- Search terms include: Functional Dyspepsia, Medicine Students, Prevalence (See Appendix 1)

### Data extraction

The data extracted from the selected articles will include:

- Study details: first, author, corresponding author, article title, country, year of publication and year of data collection
- Subjects: number of participants, range of age, population source, location, inclusion and exclusion criteria of studies selected
- Methods to assess functional dyspepsia: definition used, name of the tool, among others.
- Results: prevalence of dyspepsia in general and according to sex.

In case we find a study in which the methodology or outcome is not clearly specified, we will try to contact the corresponding author. If we do not receive an answer, then the study will be excluded from the systematic review.

### Risk of Bias and Methodological Quality of Studies

We will assess risk of bias using a tool developed and validated by Hoy et al. to assess prevalence studies. (4)

### Statistical analysis

We will conduct a narrative synthesis and summarize the information collated such as means and prevalence estimates for each of the articles or reports included. If possible, we will quantitatively summarize the information by conducting a random-effects meta-analysis to pool available metrics into one summary estimate (e.g., mean of prevalence estimate). We will also aim to conduct stratified analysis by sex.

## Data Availability

Carmona-Sanchez R. Editorial al articulo titulado Factores asociados a dispepsia no investigada en estudiantes de 4 facultades de medicina de Latinoamerica: estudio multicentrico. Revista de Gastroenterologia de Mexico. 2018;83(3):213-214.
Ford, Alexander C., et al. Global prevalence of, and risk factors for, uninvestigated dyspepsia: a meta-analysis. Gut 64.7 (2015): 1049-1057.
Vargas-Matos, Iván, et al. Superposicion del sindrome de intestino irritable y dispepsia funcional basados en criterios ROMA III en estudiantes de medicina de una universidad privada de Lima, Perú. Revista de Gastroenterologia del Peru 35.3 (2015): 219-225.
Hoy, D., Brooks, P., Woolf, A., Blyth, F., March, L., Blain, C., & Buchbinder, R. (2012). Assessing risk of bias in prevalence studies: modification of an existing tool and evidence of interrater agreement. Journal of clinical epidemiology, 65(9), 934-939.
Basandra S. Epidemiology of Dyspepsia and Irritable Bowel Syndrome (I BS) in Medical Students of Northern India. JOURNAL OF CLINICAL AND DIAGNOSTIC RESEARCH. 2014.
Jaber N, Oudah M, Kowatli A, Jibril J, Baig I, Mathew E et al. Dietary and Lifestyle Factors Associated with Dyspepsia among Pre-clinical Medical Students in Ajman, United Arab Emirates. Central Asian Journal of Global Health. 2016;5(1)
Vargas M, Talledo-Ulfe L, Samaniego RO, et al. Dispepsia funcional en estudiantes de ocho facultades de medicina peruanas. Influencia de los hábitos [Functional dyspepsia in students of eigth peruvians medical schools. Influence of the habits]. Acta Gastroenterol Latinoam. 2016;46(2):95-101.
Mejia Christian R, Quezada-Osoria Claudia, Verastegui-Diaz Araseli, Cardenas Matlin M, Garcia-Moreno Katerine M, Quinones-Laveriano Dante M. Factores psicosociales y hábitos asociados con dispepsia funcional en internos de un hospital nacional en Piura, Perú. Rev Col Gastroentero. 2016 Dec; 31(4): 354-359.
Canales-Pichen, Diego, and Javier Carhuaricra-Atahuaman. Dispepsia funcional en estudiantes de medicina de la Universidad Nacional Hermilio Valdizán, 2017. Revista Peruana de Investigación en Salud 3.1 (2019): 36-42.

https://www.sciencedirect.com/science/article/pii/S0375090618300569

https://pubmed.ncbi.nlm.nih.gov/25147201/

https://pesquisa.bvsalud.org/portal/resource/es/lil-790095

https://pubmed.ncbi.nlm.nih.gov/22742910/

https://www.ncbi.nlm.nih.gov/pmc/articles/PMC4316280/

https://www.ncbi.nlm.nih.gov/pmc/articles/PMC5661185/

https://pubmed.ncbi.nlm.nih.gov/28703562/

https://pesquisa.bvsalud.org/portal/resource/es/biblio-960031

http://revistas.unheval.edu.pe/index.php/repis/article/view/253

## APPENDIX 1 SEARCH TERMS AND STRATEGY

### Search term for Pubmed

(“Dyspepsia”[Mesh] OR dyspepsia OR heartburn OR “Rome III” OR “Rome IV” OR “Gastrointestinal Diseases”[Mesh] OR Functional gastrointestinal disorder) AND (Medical students OR medicine students OR Pre-medical students OR Pre-clinical students OR health students OR “Students, Medical”[Mesh])

### Search term for Global Index Medicus

Tw:(dyspepsia OR heartburn)) AND tw:(medical students OR health students))

### Search term for EMBASE

(‘dyspepsia’/exp OR ‘heartburn’/exp OR ‘rome iii’ OR ‘rome iv’ OR ‘digestive system function disorder’/exp) AND (‘medical student’/exp OR ‘medicine students’ OR ‘pre-medical students’ OR ‘pre-clinical students’ OR ‘health students’)

## Notes

**Conflicts of interest:** All authors declare to have no conflicts of interest.

### Competing Interest Statement

The authors have declared no competing interest.

### Funding Statement

No funding provided for this study

### Author Declarations

This is a systematic review and meta-analysis protocol. No human subjects were involved in this project. Thus, this study was classified as non-human subject research. Therefore, no approval was needed from an IRB/ethics committee

